# PROFILE AND TRAINING ON GENDER IDENTITY AMONG HEALTH PROFESSIONALS IN THE CAETÉ REGION IN THE EASTERN AMAZON

**DOI:** 10.1101/2023.04.05.23288180

**Authors:** Pedro Renan Nascimento Barbosa, Roseane do Socorro da Silva Matos Fernandes, Erica Feio Carneiro Nunes, Cibele Nazaré Câmara Rodrigues

## Abstract

**Background:** The National Policy on Integral Health of Lesbians, Gays, Bisexuals, Transvestites, and Transsexuals (PNSILGBT+) aims to eliminate discrimination and institutional prejudice in public health services, with emphasis on training and enabling professionals to assist in the care of this population. However, for transsexuals and transvestites, this access continues to be neglected.

**Aim:** To determine the profile of health professionals, their academic-professional training in health, and their knowledge about the specificities of gender identity.

**Methods:** This is a cross-sectional, observational, quantitative survey conducted with health professionals working in the Caeté Region. Data were collected using a semi-structured questionnaire, which was applied online via Google Forms in the period from March 2021 to August 2021.

**Outcomes:** Epidemiological profile, academic education, area of practice, and knowledge about gender identity were predictors for the descriptive analysis.

**Results:** The sample consisted of 73 participants: 82.19% female, 100% cisgender, with a mean age of 32.37±7.80 years, and the majority self-declared as Catholic (65.75%). The largest number of health professionals were in Nursing (34.24%), practicing in Tertiary Care (50.68%), with an average time of service of 6.24±6.30 years. The volunteers reported that they did not understand the difference between cisgender and transgender people (57.53%), admitting that they had not received any training on gender identity or the healthcare of Lesbians, Gays, Bisexuals, Transvestites, and Transsexuals (LGBT)+ people (79.45%). However, 87.67% understood the importance of training on this subject for the healthcare of this population.

**Clinical Implications:** This study demonstrates the need for continued trainings on gender identity, health and care of the LGBT+ population, with a focus on transgender people, to eliminate institutional bias.

**Strengths and limitations:** The research was based on the search for professionals’ knowledge about gender identity, even in healthcare in a region in the interior of the Amazon, however it did not reach the preterit sample n, but it was important for the debate between the professionals reached and the institutions and their managers.

**Conclusion:** The academic training and professional development of health professionals are still lacking when it comes to the theme studied; a fundamental point needed to comply with the National Curricular Guidelines (NCDs) for courses in health, National Policy for Professional Development, and in particular, the PNSILGBT+.

## INTRODUCTION

The National Policy on Integral Health of Lesbians, Gays, Bisexuals, Transvestites, and Transsexuals (PNSILGBT+) established in Ordinance 2,836 of December 1, 2011, of the Brazilian Ministry of Health, promotes integral LGBT+ health based on the elimination of discrimination and institutional prejudice in public health services. It also contributes to reducing inequalities and violations of the rights of this social minority and to consolidate the guiding principles of the Unified Health System (*Sistema Único de Saúde—*SUS) as universal, integral, and equitable for quality care (1)

The discussions that emerged around this Policy bring to light the vulnerability to which the LGBT+ population is subject, due to the non-fulfillment of their fundamental citizenship rights when it comes to healthcare. This raises questions about professional training and the care of this population within the SUS. Such debates consider that healthcare professionals need to be qualified to receive this social minority groups^1*^, whether in the practical care provided or to demystify any prejudiced conceptions; this requires a new approach to healthcare (2).

Due to the non-equivalence of gender and biological sex, the transsexual and transvestite populations are those that most frequently suffer attacks against their human and basic rights. Aware of this reality, the Ministry of Health recognizes that sexual identity and gender identity are part of a complex process of discrimination and exclusion from which factors of vulnerability, such as violation of the right to health, dignity, non-discrimination, autonomy, and free development, derive (3,4).

Composing an analytical matrix, Zerbinati (5), creates a temporality of historical accounts needed to discuss gender fluidity, starting with ancient times, with the divinity of Príapo, son of the goddess Aphrodite and the god Dionysus, in which he is characterized as androgenous and hermaphrodite, playing a decisive role in human and agricultural fertility (6); then moving forward to the French Revolution, with its ideals that still influence our understanding of female empowerment^2**^ and the discussions about the social roles of man and woman and the deconstruction of the previously established standard (7).

From the eighteenth century, marked by the French Revolution and its liberal ideals, an individual’s status (man or woman) came to be adapted according to the development of society and the process of humanistic analysis beyond the merely biological, through a process of formation of their identities, particularly women, which impacted on the self-view of their gender identity and sexual orientation (6,7).

In the nineteenth century, the importance of discussions related to human sexuality expanded to include the medical-social approach, due to the impacts of gender-related sexual disorders and their repercussions on society, work, family life, and other aspects of human life (8). However, it is only during the twentieth century, in the 1960s and 1970s, with the appearance of LGBT+ social movements, that the medical community began to intervene in the care of the LGBT+ population (6).

With the biomedical positions aligning with the process of transgender visibility and playing an active role in the process of sexual reassignment, the understanding and the need to separate sexual identities into categories has become prominent; for example, not confusing homosexuality with transsexuality and/or transvestility, but rather, focusing on its characteristics, and whether or not the sex assigned at birth should be accepted (6).

This historical context demarcates not only the urgent need to overcome the pathological/prejudiced view of transgender subjects who belong to the LGBT+ community, but the need for multi-professional healthcare, and professionals who have a knowledge and understanding of the socio-historical and cultural factors involved, to intervene in their process of transition and sexual reassignment, for example (3,4).

However, studies indicate that two out of three LGBT individuals (67%) have suffered some type of discrimination related to their sexual identity or gender, and this figure is as high as 85% among transvestites and transsexuals. The data in this Document also indicate that in a study conducted at the Gay Pride Parade in São Paulo, 14.5% of those questioned reported having suffered some kind of prejudice concerning the healthcare services received (4,9,10).

All these aspects demonstrate the need for health professionals to ensure humanized conduct and a good understanding of aspects related to the diversity of this population, its sexuality, and its specificities in interpersonal/affective relationships^3^ in healthcare. Out of this understanding by training institutions, curricula emerge that guarantee health specificities in professional practices and the proper handling of pronouns and chosen names, for example, contributing substantially to the improvement and quality of access to health services (3,4).

Because of the above, this research aims to verify the academic-professional training profile in healthcare, and their knowledge about the specificities of gender identity.

## METHODS

This is a quantitative, observational, cross-sectional study. The participant profile was defined as health professionals in the Caeté Region (Eastern Amazon), in professional practice at any level of healthcare (Primary Care, Secondary Care, Tertiary Care, or Managerial). The research was submitted to the Research Ethics Committee of the Center for Tropical Medicine of the Federal University of Pará (*Núcleo de Medicina Tropical da Universidade Federal do Pará—*CEP*—*NMT/UFPa: CAAE— 40179820.6.0000.5172) and was approved through Opinion No. 4,514,620.

The participants were contacted via e-mail through data provided by the Municipal Health Departments of the National Registry of Health Establishments (*Cadastro Nacional de Estabelecimentos de Saúde—*CNES). The participants were informed about the research through an Informed Consent Form (ICF), guaranteeing their freedom to participate voluntarily, and informing them that they were free to discontinue at any stage of the study, besides the questionnaire being totally confidential, with no personal identification.

For the collection of empirical data, a semi-structured questionnaire was used, with 23 questions. The questionnaire was applied online using Google Forms. The first part aimed to gather the participants’ profiles in terms of socioreligious and demographic aspects, and professional education (age, sex, gender, religion, religious practice, educational institution, training area, and level of healthcare practice, among others) and the second part contained fundamental questions on the theme (gender identity, the difference between cisgender and transgender, sexual organ designated at birth for each gender identity, chosen names, academic training and professional development on the theme, and need for training).

The survey was conducted between March 2021 and August 2021. A total of 267 questionnaires were distributed, giving a deadline of up to 48 hours for their return. Of the 103 questionnaires that were returned, 30 were excluded because they had incomplete responses and/or did not meet the inclusion criteria of the study. The quantitative data generated from the objective questions were transferred from Google Forms to Excel Office 2019, and tabulated and organized for analysis and discussion (see tables below).

## RESULTS

The sample consisted of 73 participants who met the established inclusion criteria and agreed to participate by signing the ICF. Of these, 82.19% (n=60) were female and 17.80% (n=13) male; 100% of the sample identified as cisgender, with a mean age of 32.37 years (Standard Deviation [SD] = 7.80). As regards religion, the majority declared themselves Catholic (65.75%) and practicing in their faith (58.90%). The data are shown in Table 1.

**Table 1.** Socioreligious and demographic data (n=73)

|  | n | % |
| --- | --- | --- |
| Sex |  |  |
| Female | 60 | 82.19 |
| Male | 13 | 17.80 |
| Gender Identity |  |  |
| Cisgender man | 60 | 82.19 |
| Cisgender woman | 13 | 17.80 |
| Other | 0 | 0 |
| Age group |  |  |
| 21-25 | 24 | 32.87 |
| 26-30 | 13 | 17.80 |
| 31-49 | 34 | 46.57 |
| ≥ 50 | 2 | 2.73 |
| Religion |  |  |
| Catholic | 48 | 65.75 |
| Evangelical | 16 | 21.91 |
| Other | 2 | 2.73 |
| No religion | 7 | 9.58 |
| Practicing |  |  |
| Yes | 43 | 58.90 |
| No | 23 | 31.50 |
| No religion | 7 | 9.58 |

In the reality studied, the largest number of health professionals in the sample worked in the area of Nursing, with 34.24% (n=25), followed by Physiotherapy, with 13.69% (n=10) and Psychology, with 13.69% (n=10).

Regarding the level of healthcare, a majority of the participants worked in Tertiary Care (50.68%); notably, 26.02% (n=19) indicated that they worked in two or three levels of healthcare or management. The average length of service was 6.24 years (SD = 6.30). The data are shown in Table 2.

**Table 2.** Professional training, level of healthcare, and length of service (n=73)

|  | n | % |
| --- | --- | --- |
| Training |  |  |
| Biomedicine | 5 | 6.84 |
| Nursing | 25 | 34.24 |
| Physiotherapy | 10 | 13.69 |
| Pharmacy | 4 | 5.47 |
| Speech therapy | 5 | 6.84 |
| Medicine | 4 | 5.47 |
| Nutrition | 2 | 2.73 |
| Psychology | 10 | 13.69 |
| Social services | 6 | 8.21 |
| Occupational therapy | 2 | 2.73 |
| Level of Attention in action |  |  |
| Basic | 21 | 28.76 |
| Secondary | 29 | 39.72 |
| Tertiary | 37 | 50.68 |
| Management | 8 | 10.95 |
| Time of Professional Performance |  |  |
| > 1 year | 12 | 16.43 |
| 1–5 years | 31 | 42.46 |
| 6–10 years | 18 | 24.65 |
| < 10 years | 12 | 16.43 |

When questioned about their knowledge of gender identity or care for LGBT+ people, 57.53% of the participants demonstrated that they did not know the difference between cisgender and transgender (n=40); however, 69 individuals (94.52%) understand what is meant by a preferred, or chosen name.

When asked whether they had received any training in this area, the majority (79.45%) admitted to not having received any training on gender identity and/or healthcare of LGBT+ people; however, 64 of the participants (87.67%) understood the need for training on this theme for their routine in healthcare, as shown in Table 3.

**Table 3.** Information on Professionals’ Knowledge About Gender Identity and/or Healthcare of LGBT+ people (n=73)

|  | n | % |
| --- | --- | --- |
| Do you know the difference between transgender and cisgender? |  |  |
| Yes | 31 | 42.46 |
| No | 42 | 57.53 |
| Do you know what a chosen or preferred name is? |  |  |
| Yes | 69 | 94.52 |
| No | 04 | 5.47 |
| Have you received any support training in gender identity and/or healthcare for LGBT+ people? |  |  |
| Yes | 15 | 20.54 |
| No | 58 | 79.45 |
| Do you believe it is important to have training on gender identity and/or healthcare for LGBT+ people? |  |  |
| Yes | 64 | 87,67 |
| No | 9 | 12,32 |

## DISCUSSION

When discussing gender identity, it is important to emphasize that “gender” is related to traits/characteristics of socio-psycho-cultural origin, such as femininity and masculinity, and to the notion of social groups of what is considered feminine or masculine; it is also linked to stereotypes^4*^ and/or what society expects and attributes to what it is to be a man or to be a woman (11). This is different from “sex,” which relates purely to the biological aspects: reproductive anatomy, chromosomal formation of the genome, gonads/hormones, and sexual behavior, according to traits and characteristics of biological etiology (12).

Thus, we see a contradiction between what it is to be a man or a woman, going beyond biological discussions and entering the sociocultural sphere; the result of the construction of the environment in which a person lives, reaffirming the words of Simone de Beauvoir (13): “*On ne naît pas femme, on devient femme*”^5**^, which reflects the meaning of the construction of being a woman and the social, cultural, and political impacts contained in this condition; aspects that should be considered in the identity process, moving away from the idea that associates all this dynamic of “being” with the characteristics merely of biological etiology (14).

It should be mentioned that gender identity^6***^ is on the agenda of discussion in contemporary society when it comes to issues related to transsexuality or transvestility, or even non-binarity, in which both break with the culturally standardized cisgender concept from ancient times, with the biblical figures and based on the origins of humankind, according to Creationism^7****^, as Scott (15) points out in his historical analysis of gender construction.

Medicine and other sciences interested in discovering the foundations of the human body needed to create definitions of sexual practices and identities(16). Thus, from the twentieth century, a process of biomedical deconstruction began to take place, seeking to understand the phenomenon of health/sickness as it relates to the transgender public, going beyond the term “transsexualism,” which refers to the pathology provided for in the International Classification of Diseases—ICD 10 (17,18), code F640, which defines transsexuals as individuals “affected by disease, capable of undergoing reparative surgical interventions, able to adapt their physical body to the sex they have in their mental representation” (19,20).

From this nomenclature and all the prejudiced analysis that surrounded and involved it, “transsexualism” configured as “gender identity disorder” was later replaced by “gender identity disorder” and the current Diagnostic and Statistical Manual of Mental Disorders – DSM-5 (21–23), it is called “gender dysphoria,” which, unlike the previous classifications, views transsexuality as a “psychological state of acute suffering that requires intervention, above all, medical” (16).

These milestones in the scientific literature on health and diagnosis involve the urgent need for professionals who act in the transsexualizing process, with the panorama and training demystified concerning the factors involved in perpetuating the prejudiced and pathological view of gender dysphoria, composing multidisciplinary and interdisciplinary teams that understand this process and in the specificities of the individual. However, the data shown in Table 3 demonstrate this unpreparedness, as 42 participants (57.53%) were unaware of basic concepts of gender identity (3.4).

In this discussion, it is important to question Brazilian religiosity, which influences the national scenario in various ways; it acts as a positioning (ideology) and as an instrument of social intervention, intervening in the day-to-day political decisions (repressing or releasing some practices) and in judicial policy and state decisions. Therefore, in the current social context, in which various religious leaders are also present in the legislative and executive spheres of the country, they influence their ideologies with regard to decision-making on issues of relevance to these citizens (24).

Since religion is important in social configurations, its convictions, its positions, and its actions in the field of sexuality influence the experiences and policies of sexuality and gender, being prioritized by religious leaders, especially from a Judeo-Christian perspective, as a matter of debate and negative regulation, gender themes, such as abortion and discussions about the LGBT+ population, go against the predictive concept of the Secular State^8*^, to which Brazil belongs (24–26).

In Jewish-Christian religious traditions, the sexual behavior of LGBT+ people demonstrates spiritual weakness to resist “demonic temptations,” in violation of biblical beginnings, and moving away from the divine and His plan for families, idealized by the figures of Adam (a heterosexual cisgender man) and Eve (a heterosexual cisgender woman), created in the formation of humanity, according to Genesis (27,28).

Moreover, from the religious perspective, Fernandez (29) shows the influence of healthcare professionals’ religion on the care provided, with worrying reports that contradict the perspective desired by the SUS, with its policies of comprehensive, equitable, and universal health for every citizen, especially for LGBT+ users, corroborating Roncon (30) and Souza (31.32), who emphasize discriminatory practices, through religious discourses on transgender people, generating mental anguish due to ignorance or neglect of scientific foundations that break with the demonic idealization about these communities, and the social prejudice that surrounds this portion of the population^9*^.

Psychological follow-up, guidance on the correct use of hormonal medications, regular consultations with the health team, evaluations for surgical interventions, performing surgeries, and the post-surgical rehabilitation process are some of the demands to be met, as many LGBT+ subjects have been on the waiting list for a long time (sometimes years). Therefore, the health team must ensure humanized and unprejudiced conduct (4).

These are characteristics that the health professional should demonstrate in the care of the LGBT+ public, specific people with gender dysphoria, since the data from this research indicates a lack of knowledge about gender identity and a lack of training on this theme (Table 3), which prompts two questions related to the educational processes: does academic education (initial education) fail to approach/address gender themes in the curricula of health professionals? How does the professional development of health professionals take place, or how should it take place, in relation to this subject and the public?

Visgueira (33), analyzing the knowledge of medical students, for example, about gender identity (n=122), showed high percentages of inadequate responses regarding the differences between definitions and specific behaviors of the transsexual population; moreover, the approach to the subject during the undergraduate course was minimal or non-existent; data that corroborate the analysis in this study, which showed that 58 of the participants (79.45%) reported not having received any training on the theme of LGBT+ and gender identity.

This panorama also reflects the inefficiency of Higher Education Institutions (HEIs) with regard to gender discussions, how these relate to the health–disease process, and their approaches to the care of the LGBT+ population, with this being an important requirement for compliance with the National Curricular Guidelines (NCDs) for health professionals, which define, as part of the professional profile, an academic background with a generalist, humanist, critical, and reflexive basis in ethical principles and understanding of social, cultural, and economic reality, acting for the benefit of society, at all levels of healthcare, based on scientific and intellectual rigor (34,35).

Another finding of this study is that of the 37 participants aged between 21 and 30 years (50.67%), 34 had a service experience of 5 years or less (94.59%; n=37). Therefore, they were mostly new graduates, corroborating the concept that there is a lack of focus on LGBT+ health in the curricula. It should be noted, however, that although the principles recommended in the conduct of health professionals are established in these NCDs, which extend to any citizen who needs care and follow-up, the data and concerns of this study justify the criticisms made here, especially those related to access to health services for transsexual people (34,36,37).

It is understood that there is a need for more training, given that most of the participants (87.67%) said they hoped to achieve the characteristics and perceptions that professionals need to have to serve the LGBT+ population (38.39); understanding gender dysphoria and the specificities of care for this public, in compliance with the fundamental principles of the SUS: health as a right of all (universality), applying prevention, promotion and rehabilitation actions to health (integrality) and reducing inequalities by offering the necessary support to those who need it most (equity) (40– 42).

The lack of debate about the care of this portion of the population during academic training becomes critical to fulfilling the National Policy on Continuing Education of the Brazilian Ministry of Health (43) for the development of the professional in service, training him/her for necessary actions in the areas in which they work, finding solutions to the problems they face, articulating professional practice and learning, and effectively implement public health policies such as the PNSILGBT+ (44.45).

With these specificities in the care and treatment of the transgender public and members of the LGBT+ community, in 2007, during the 13^th^ National Health Conference (CNS), through the implementation of the PNSILGBT+, approved only in 2009 and instituted in the SUS in 2011, measures were taken to guarantee sexual and reproductive rights, professional development practices in health, and review of the academic curricular, encouraging the production of scientific research, technological innovation and the sharing of therapeutic advances, protocols to combat violence, non-discriminatory regulation of blood donations, the adaptation of forms and medical records and the health information system, to bring into its texts, for the first time, the distinction between gender identity and sexual orientation (46).

Another milestone was reached at the 1st National Conference of Lesbians, Gays, Transvestites, and Transsexuals, held in Brasília/DF, in 2008; based on the Federal Constitution of 1988, strategic guidelines were created aimed at promoting citizenship and human rights to meet the specificities of lesbian, gay, bisexual, transvestite, and transsexual scare in the health–disease process (2).

In that same year, GM Decree 1707, of August 18, 2008, which established the transsexualizing process under the SUS, and SAS Decree of August 19, 2008, which approved and regulated this process in the SUS, was forwarded to public consultation, in the period from June 26 to July 30, 2008, installed in the DATASUS consultation tool, and published in the DOU through Ordinance 1279, of July 25, 2008 (2.46).

The National Policy for Integral Healthcare for Men (*Política Nacional de Atenção Integral à Saúde do Homem*)(47), instituted in 2009 by the Ministry of Health within the SUS, through Decree 1,944 of August 27, 2009 (48), guarantees the specificities of gay, bisexual, transvestite, and transsexual men, with the publication of its version of the Basic Care Notebook no. 26, Sexual and Reproductive Health (*Caderno da Atenção Básica n*^*0*^ *26, Saúde Sexual e Reprodutiva*), which deals with the themes of sexual orientation and gender identity, aimed at providing guidance and training for health professionals to care for the LGBT+ population (49).

The abovementioned policies initiated the transformation of the SUS as the main sector in the care and assistance provided to transgender people. This research, with its participants and its data, aims to reinforce the adaptations in care and assistance of this public, encouraging debates at the curricular level among future professionals, and as part of the professional development of professionals, among other aspects needed to expand the knowledge of working professionals, and break the cycle of institutional and professional prejudice that pose barriers to the inclusion and adaptation of transvestites and transsexuals when it comes to health services.

During this research, there were limitations in terms of the completion of questionnaires by the participants. The questionnaires were sent out online, at the recommendation of the Research Ethics Committee, due to the issues faced worldwide by the COVID-19 Pandemic; there were also difficulties in accessing the questionnaire, and a fear among the participants of getting the questions wrong, and these impressions may have led to them withdrawing their participation in the survey, or failing to complete the questionnaires.

## FINAL CONSIDERATIONS

The PNSILGBT is a milestone in the fulfillment of the human rights of transsexual and transvestite people when it comes to access to public health services and the effective treatment and follow-up by the multidisciplinary team, as part of the transition and sexual reassignment process. However, there is still a knowledge of the specificities that concern these processes among the professionals themselves, which hinders the dissemination of discrimination and institutional prejudice, causing this policy to be less effective.

The data from this research prompts debate and reveals gaps in professional health education for the care of the LGBT+ population, whether as part of the initial academic training or as part of ongoing professional development. This discussion rests in the NCDs of courses in health, in the absence or lack of professional development on the subject.

Given these questions, it is recommended that HEIs, health facilities, health teams, their managers, and professionals seek to understand this information and the repercussions on healthcare for the LGBT+ population, especially transvestites and transsexuals, to strengthen the outworking of the policies highlighted in this study, such as the characteristics recognized in it for the professional profile needed to work in the SUS and establish a commitment to its guiding principles.

## Data Availability

We declare Data Availability

## Acknowledgment

to the Santo Antônio Maria Zaccaria Hospital (HSAMZ), the City Hall and Municipal Health Department (SEMUSB) of Bragança, the Ministry of Education and the Ministry of Health, the Coordination for the Improvement of Higher Education Personnel (CAPES) and the Federal University of Pará (UFPa) through the Multiprofessional Residency in Women’s and Children’s Health and the Dean’s Office for Research and Graduate Studies, which funded this publication through the Qualified Publication Support Program (PAPQ).

## Footnotes

1* Refers to a human or social group that is in a situation of inferiority or subordination in relation to the other, which is considered the majority or dominant group due to various factors, such as socioeconomic, legislative, psychic, age-related, physical, linguistic, gender-related, ethnic or religious (50).

2** This is the multidimensional process of transformation of the social role of women, from subordinate to the opposite sex to an active agent, capable and with individual and collective decision-making power (51)

3 This is the process of acquisition of human and social skills needed for professional development; carrying out the services; teamwork, didactics, and reflection on the values (52)

4* Exaggerated belief associated with a category.

5** “One is not born, but becomes a woman” (13)

6*** Term used from the end of the 20^th^ century (15).

7*** The Christian belief that God created the world and all living beings, as described in the Biblical book of Genesis.

8* Promotion of the segmentation of State and Religions, not granting mediation of leaderships, streams of thought, and religious ideals in state matters, or favoring one or several religions over the others (53). ^9*^ Type of discrimination related to public or private institutions that reinforce the exclusion and marginalization of social minorities (54).

